# Arterial hypertension and its covariates among nomadic Raute hunter-gatherers of Western Nepal: a mixed-method study

**DOI:** 10.1101/2022.07.28.22277961

**Authors:** Tapendra Koirala, BC Udaya Bahadur, Carmina Shrestha, Ujjawal Paudel, Rolina Dhital, Sunil Pokharel, Madhusudan Subedi

**Author notes:** **Corresponding author** Tapendra Koirala, ^1^ Dasharathpur Primary Health Care Center, Surkhet, Karnali Province, Nepal.

## Abstract

**Objectives:** To determine the prevalence of, and understand the factors associated with, hypertension among the nomadic Raute hunter-gatherers of Western Nepal.

**Design:** A mixed-method study.

**Setting:** The study was carried out at Raute temporary campsites in the Surkhet District of Karnali Province between May to September 2021.

**Participants:** The questionnaire-based survey included all males and non-pregnant females of the nomadic Raute group aged 15 years and above. In-depth interviews were conducted among purposively selected 15 Raute participants and four non-Raute key informants to help explain and enrich the quantitative findings.

**Outcome measures:** The prevalence of hypertension (defined as brachial artery blood pressure of systolic _≥_140 mm Hg and/or diastolic _≥_90 mm Hg) and its socio-demographic, anthropometric, and behavioral covariates.

**Results:** Of the 85 eligible participants, 81 [median age 35 years (interquartile range: 26–51), 46.9% female] were included in the final analysis. Hypertension was found in 10.5% of females, 48.8% of men, and 30.9% of the total population. Current alcohol and tobacco use were high (91.4% and 70.4%, respectively), with concerning high rates among youths. Males, older people, current drinkers, and current tobacco users were more likely to have hypertension. Our qualitative analysis suggests that the traditional forest-based Raute economy is gradually transitioning into a cash-based one that heavily relies on government incentives. Consumption of commercial foods, drinks, and tobacco products is increasing as their market involvement grows.

**Conclusion:** This study found a high burden of hypertension, alcohol, and tobacco use among nomadic Raute hunter-gatherers facing socioeconomic and dietary transitions. Further research is needed to assess the long-term impact of these changes on their health. This study is expected to help appraise concerned policymakers of an emerging health concern and formulate context-specific and culturally sensitive interventions to limit hypertension-related morbidities and mortalities in this endangered population.

**Strengths and limitations of this study:**

- This is the first study to report the prevalence of hypertension and its covariates among the nomadic Raute hunter-gatherers of Nepal.
- The major strengths of this study are the use of a mixed-method design to have both quantitative and qualitative perspectives, near total population enrollment, and robust methodology.
- The cross-sectional design of this study limits its ability to establish causal relationships between the variables.
- Several important factors, such as dietary fruits and vegetable consumption, salt intake, and level of physical activities, as well as the presence of diabetes mellitus, dyslipidemia, or central obesity, were not assessed, preventing this study from determining the community’s actual cardiovascular disease risk.
- Interviews taken in language non-native (Nepali) to the Raute may be subject to language bias.

## Introduction

Hunting-gathering is one of the oldest modes of subsistence, where most or all food is obtained through foraging edible plants and hunting wild animals.[1] Hunter-gatherers (HGs) were estimated to account for about 1 percent of the world’s population in the 1960s.[2] As a result of rapid population growth, transformations of habitat, and globalization, the population of traditional HG communities have rapidly declined.[3,4] Currently, only a handful of communities are considered “pure HGs”, while many have transitioned either to agriculture, pastoralism, or to a mixed economy where other adaptive strategies supplement foraging.[5,6] HGs and other small-scale populations living traditionally are found to have a low prevalence of type 2 diabetes mellitus, hypertension, and obesity.[7,8] Their remarkable cardiovascular health is often linked to their high level of physical activity; low glycemic index diet, rich in fibers and micronutrients; and limited access to processed and high caloric modern foods.[9,10] Evidence has shown the emergence of obesity, diabetes, atherosclerosis, and other lifestyle-related diseases among the former HGs as they transitioned from traditional to Western lifestyles.[11,12] Health research in HGs has much to offer to modern medicine and public health. Knowledge of the interactions between health and lifestyle, diet, and health behavior among diverse traditional populations can aid in our understanding of several non-communicable diseases (NCDs) that plague modern societies. Inspired by HG research, several studies, including randomized controlled trials, have studied the role of HG or paleolithic diets in various NCDs, from obesity, hypertension, diabetes, and dyslipidemia to multiple sclerosis among the general population.[13]

However, HG’s health has not been extensively studied worldwide, and there is still a dearth of data on the impact of HG’s lifestyle choices on their health, particularly their cardiovascular and metabolic health. Studies from the mid-to-late 20th century constitute a sizable portion of the existing research on HG’s health.[8,9,11,14–16] Additionally, most studies focus disproportionately on HG societies in South America and Sub-Saharan Africa. Asians, South Asian HG societies, in particular, are overtly underrepresented in research scrutiny. Only a handful of studies have explored the health and well-being of South Asian HGs. Data on the cardiovascular and metabolic health of contemporary HG populations in this entire region is almost non-existent, and Nepal is no exception.

Cardiovascular disease (CVD) is among the leading cause of preventable death globally. Raised blood pressure (BP) or hypertension is one of the strongest modifiable risk factors for CVDs. Approximately 34% of men and 32% of women aged 30–79 years worldwide, as well as 30% of men and 20% of women aged 15–69 years in Nepal, were estimated to have hypertension in 2019.[17,18] Nevertheless, despite being the most representative of the available large-scale data, estimates from current national-level public health surveys are said to be significantly under-representative of ethnic minorities and difficult-to-reach mobile and migrant sub-populations.[19,20] Furthermore, due to their unique lifestyle, diet, and behavior-related risks, the nationally representative data cannot be generalized to special sub-populations/groups.

Here, we present a distinctive group of Nepalese nomadic hunter-gatherers with a unique set of lifestyle, tradition, language, and socio-cultural values who continue to live their traditional life, migrating from one place to another regularly gathering wild foods and bartering their carved woodenware for the grains and other necessities from the settled villagers. The Rautes, a population apparently on the verge of extinction, is Nepal’s last remaining nomadic hunter-gatherer. Since their existence was first documented in 1955, the Raute ethnography has been extensively studied by scholars worldwide. However, the Rautes’ health remains scientifically unexplored, especially their cardiovascular and metabolic health. Although anecdotal evidence suggests socio-economic transition, changing health behaviors, and deterioration in the general health of the Rautes, no scientific studies have explored the magnitude of such change and their impact on their health. Therefore, in an attempt to understand the state of their cardiovascular health, this study aims to determine the prevalence of hypertension and understand the socio-demographic, anthropometric, and behavioral factors associated with hypertension among nomadic Raute hunter-gatherers of Western Nepal.

## Methods

### Study design

This study adopted an explanatory sequential mixed methods design. A quantitative study designed to determine the prevalence and factors associated with hypertension was followed by a qualitative study to understand, explain and enrich the quantitative findings.

### Study population

The Raute is a small group of highly migratory, egalitarian hunter-gatherers that exclusively hunts Rhesus and Langur monkeys, forages edible roots, fruits, and herbs, carves woodenwares, and trades them with the settled villagers for grains, clothes, and other necessities.[16,21] Rautes strongly opposes any idea of permanent settlement, agricultural practices, and formal education. They are uninterested in new technologies and sophisticated gadgets and have little desire to save or store items.[22] The Raute primarily inhabits forested areas and riverbanks of the mid-western part of the country. They alternate their habitat between higher altitudes (6000-10000 ft) in monsoon-summer months (April to September) and lower altitudes (2000-4500 ft) in winter (October to March).[16,22,23] Their customary periodic migration is a complex function of their socio-cultural belief system, environmental conditions, availability of forest resources, prospects for trade, and relations with locals.[16,22,24] ‘Khamchi’, a Tibeto-Burman language, is their mother tongue, although they are fluent in the local Nepali language.[23] Their current economy is based on forest resources, trade, and state incentives.[22]

### Study site and setting

Karnali province, the main home of the nomadic Rautes, is the largest but least populated of all the seven provinces of Nepal, with a population of about 1.5 million, a literacy rate of 62.77%, and a human developmental index score of 0.427. The province comprises ten districts, Surkhet being the provincial capital.[25] Districts such as Kalikot, Dailekh, Surkhet, Salyan, and Jajarkot are among the most frequently, although not exclusively, inhabited places by the Rautes. At the time of the commencement of this study, the population had been migrating through the District Surkhet. The study was completed in two phases between May to September 2021, following the Raute through their six successive campsites within the territories of Lekbeshi and Gurbhakot Municipalities of the District Surkhet (Figure 1).

**Figure 1.**
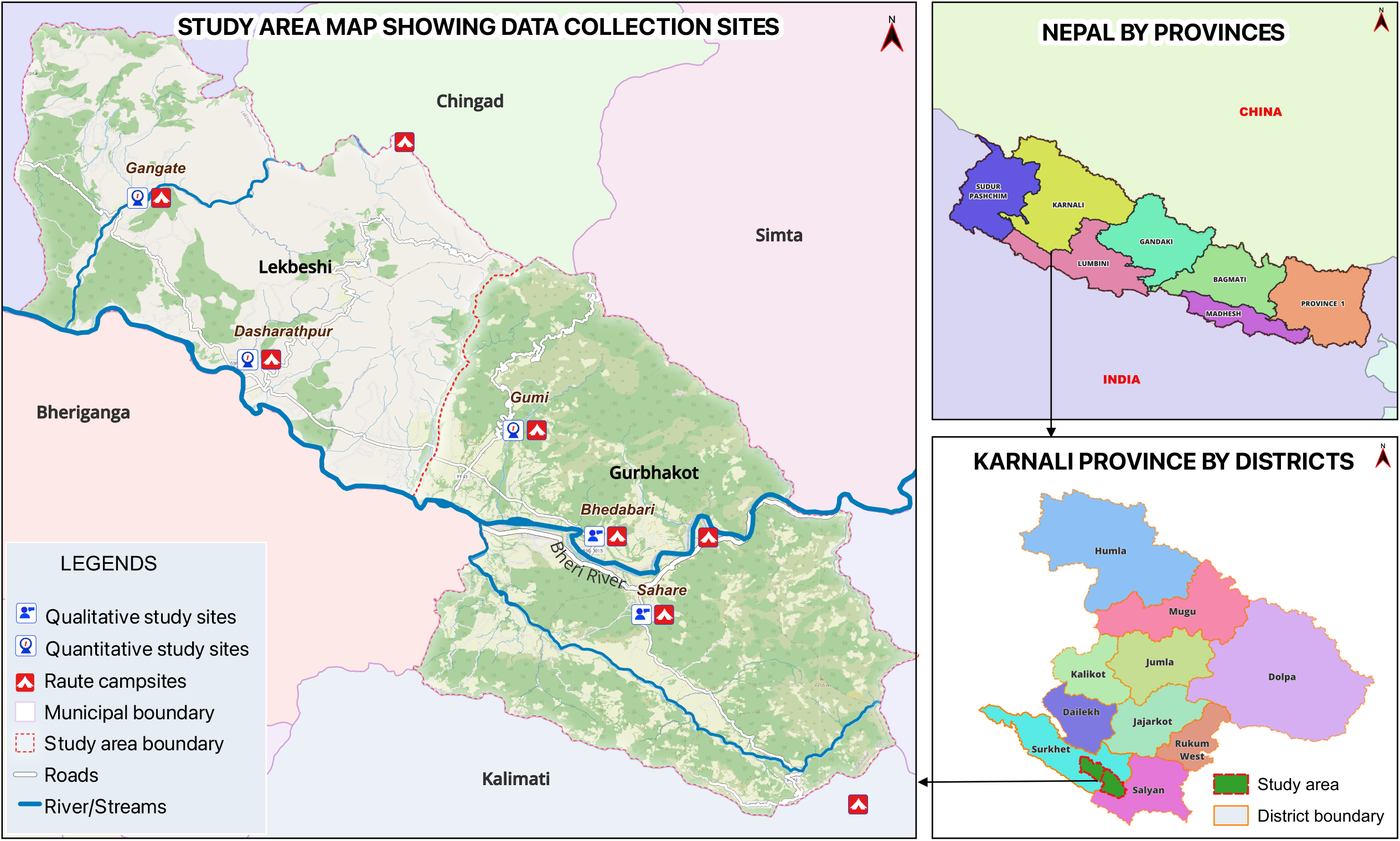
Study area map showing temporary Raute settlements (camps) and the data collection sites. Maps were created exclusively for the study using the QGIS version 3.28.0, and the GPS coordinates were taken from the study sites. (Data source: Shapefiles, Survey Department, Government of Nepal, http://nationalgeoportal.gov.np; Basemap, OpenStreetMaps contributors, https://www.openstreetmap.org)

### Participants selection

For the quantitative study, all willing individuals aged 15 years and above belonging to the nomadic Raute population were considered eligible. Participants with suspected COVID-19 and pregnant females were excluded from the study (Supplemental Figure 1). Since the entire eligible population was included, the sampling was deemed unnecessary.

Participants for the qualitative study were purposively selected: In-depth interviews (IDIs) were conducted among 15 (10 males and five females) Raute adults with and without hypertension. Four key informant interviews (KIIs) were conducted among a healthcare worker, two social workers, and an expert in the field. The participants for the interviews were selected so that each would bring a unique perspective, thereby maximizing demographic and experiential heterogeneity within the group. Interviews were conducted among Raute participants and outsiders to achieve a balanced perspective. We stopped taking further interviews when data obtained from respondents was repetitive or no new information was forthcoming.

### Data Collection

Quantitative data were collected through a structured questionnaire which included: face-to-face interviews, anthropometric measurements, and clinical examinations. Four health workers (2 Health assistants and 2 Public health nurses) were recruited and trained for data collection. They were supervised by the principal investigator (TK) and co-investigators (CS & UP) during data collection. We used standard methods to obtain quantitative measurements.[26] The data was collected at participants’ residences at their convenience by the data enumerators of their respective sexes.

Qualitative data collected through interviews (IDIs and KIIs) were taken face-to-face by two interviewers, UBB (MA) and TK (MBBS), using interview guides. UBB is a male public health expert with more than ten years of experience. TK is a male medical graduate with over five years of clinical and research experience. Interviewers had a keen interest in non-communicable diseases and their risk factors. They didn’t have any obvious bias or assumptions toward study participants. A brief informal discussion was carried out with each participant to establish rapport and facilitate participation. Every participant was informed about the aims of the study, the researcher’s personal goal, and the reason for conducting this study. All the participants approached for the study consented to participate, and there was no drop-out. Each of the IDIs lasted for approximately an hour and was audio-recorded. No repeat interviews were conducted with any participants. The interviewers made field notes during the data collection.

All the interviews, measurements, and clinical examinations were carried out, taking necessary precautions against COVID-19 transmission. The data enumerators and interviewers were tested for COVID-19 before and after carrying out data collection. During data collection, all researchers wore masks, face shields, and gloves, while all the participants were given masks and kept at a distance of at least one meter unless required to do otherwise.

### Tools and instruments

#### Survey tool

To collect quantitative data, a structured questionnaire was adapted from validated tools for various national-level surveys and other relevant literature.[18,27,28] The questionnaire was tailored according to the local context (Supplemental File 1). The questionnaire collected information on participant’s socio-demographic characteristics (age, sex, marital status, educational status, and primary occupation), health-related behaviors (alcohol consumption and tobacco use), awareness and treatment of hypertension, physical measurements (height, weight, body mass index (BMI) and BP). Physical measurements were taken by trained enumerators according to established guidelines.[26] Bodyweight in kilograms was measured to the nearest 0.2 kg using a well-calibrated standardized digital weighing scale (Seca, Hangzhou, China).

Height was recorded to the nearest 0.1 cm using well-calibrated standardized portable stadiometers (Seca, Hangzhou, China). Weight and height measurements were performed without shoes, headgear, or heavy clothing. BMI was calculated as weight in kilograms (kg) divided by height in meters squared (m^2^). BP measurements were performed using a well-calibrated digital automatic BP monitor (OMRON M6, Japan) with a universal cuff after the participants rested quietly for 15 minutes with their legs uncrossed. Each of the three BP readings was taken with participants requested to rest for three minutes in between. The mean of the second and third readings was taken for further analysis.

#### Interview guides

To collect qualitative data, we developed guides for IDIs and KIIs based on themes identified in the quantitative study. We also explored additional themes not captured in the quantitative study (Supplemental File 2).

#### Validation of tools

The survey questionnaire and interview guides were designed in English and translated into Nepali by the experts. The tools were then back-translated into English by a panel that speaks both languages for validation before administration. The structured survey questionnaire was pretested among 8 Raute participants (10% of the total sample size) who were excluded from the final analysis. Similarly, the interview guides for IDI and KII were also pretested among 3 Raute participants and one key informant before their administration.

### Operational Definitions

Operational definitions were adopted for the key variables to maintain uniformity and consistency. Hypertension was defined and categorized based on the JNC 7 recommendations.[29] Details on operational definitions of the other study variables are given in the online Supplemental file 3.

### Data management and analysis

#### Quantitative data

We used descriptive statistics such as median and interquartile range (IQR) for the continuous numerical variables and frequencies and proportions for the categorical variables to summarize our findings. All the data were analyzed in SPSS (IBM) version 23 and visualized on R version 4.1.3. Maps were created by using QGIS version 3.28.0.

#### Qualitative data

For qualitative analysis, the audio recordings of IDIs and KIIs were transcribed verbatim in Nepali and then translated into English by the two investigators, TK and CS, independently. All the transcripts were imported to the Dedoose version 8.2.14 (SocioCultural Research Consultants, LLC, Los Angeles, CA). Two investigators, TK and RD, independently reviewed and coded the transcripts. Codes were compared, and the disagreements were resolved by consensus. All codes were merged into similar categories, which were further developed into major themes inductively. The data acquired through KIIs, IDIs, and the quantitative study was triangulated during the final analysis. We did not return the transcripts to the participants due to logistical difficulties in finding highly migratory participants and their low literacy.

#### Data management

Participants’ personal health information (PHI) was protected on researchers’ fully encrypted devices with procedures for the deidentification of data during analysis, and no PHI was linked to geospatial data in the public domain. To ensure the credibility of the analysis, all procedures were supervised by research experts in the field.

### Patient and public involvement

Raute participants were included during pretesting of quantitative and qualitative tools. Their feedback was incorporated accordingly. Raute chieftain coordinated with researchers to help them gather the necessary data. Social workers and relevant public health authorities will help disseminate the results to the participants.

## Results

### Quantitative results

#### Population characteristics

Of the total 85 eligible participants, 81 participants between 15 to 69 years of age were included in the final quantitative analysis (Supplemental Figure 1). The non-response rate was 2.4%. Table 1 Summarizes the key population characteristics of this study. The median age of the participants was 35 years (IQR: 26–51), with a slight male predominance (53.1%, n=43). The majority of the participants were currently married (64.2%). Most males (90.7%) reported ‘carving and trading woodenware’ as their main occupation, while the majority (94.7%) of females reported being homemakers. None of the participants had received any formal education.

**Table 1.**
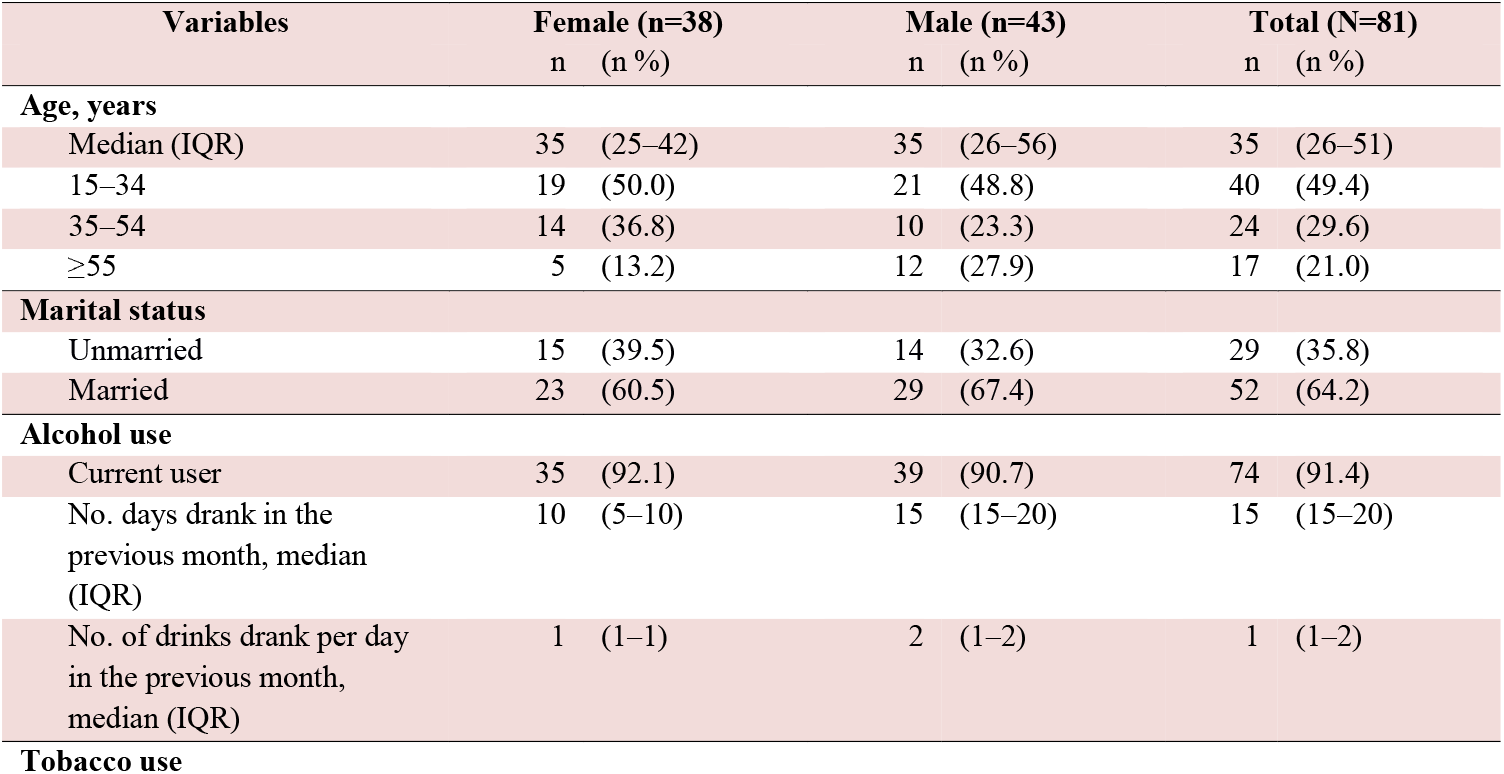
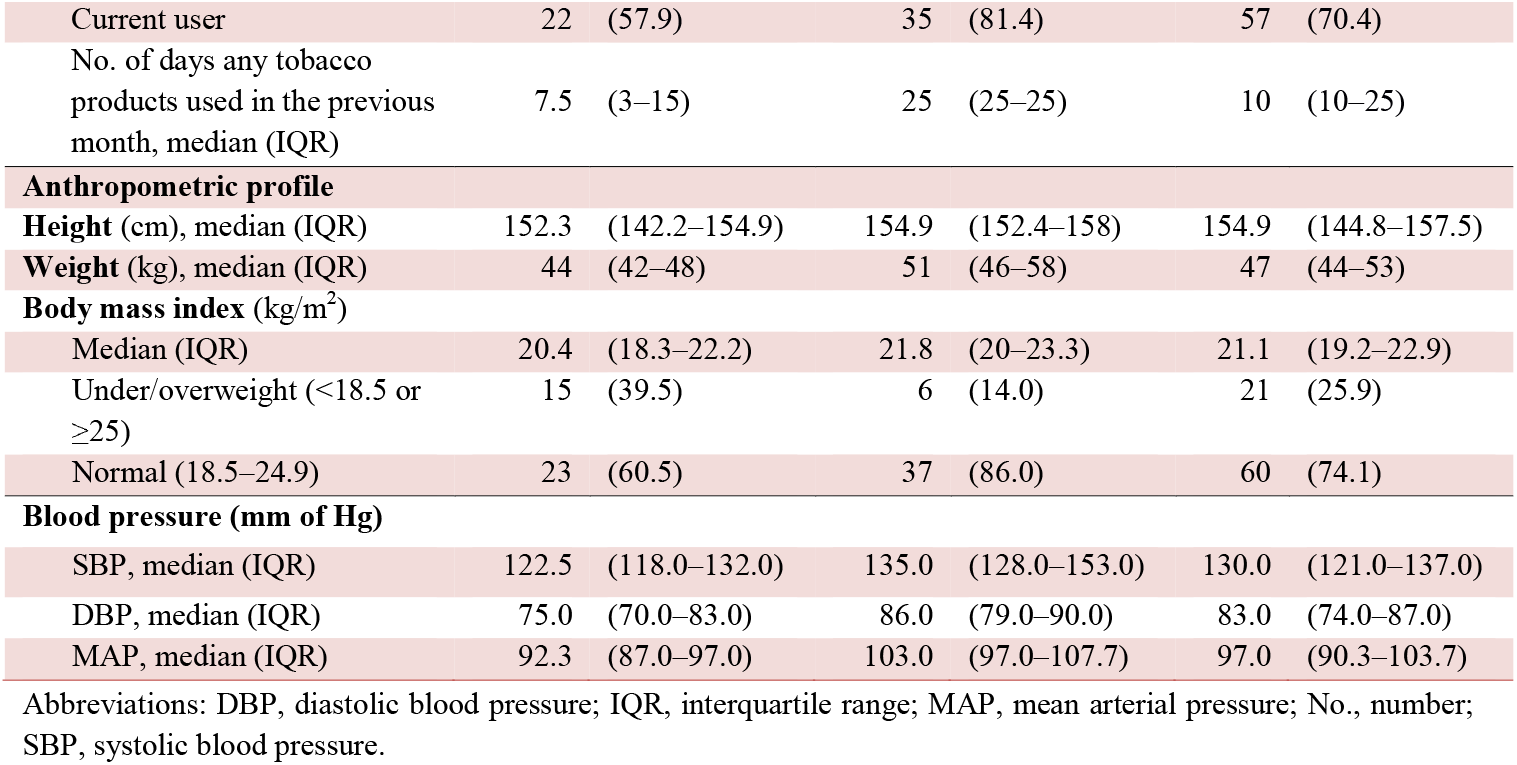
Socio-demographic characteristics, behavioral risk factors, anthropometric profile, and blood pressure of the nomadic Raute population by sex

| Variables | Female (n=38) |  | Male (n=43) |  | Total (N=81) |  |
| --- | --- | --- | --- | --- | --- | --- |
|  | n | (n %) | n | (n %) | n | (n %) |
| **Age, years** |  |  |  |  |  |  |
| Median (IQR) | 35 | (25–42) | 35 | (26–56) | 35 | (26–51) |
| 15–34 | 19 | (50.0) | 21 | (48.8) | 40 | (49.4) |
| 35–54 | 14 | (36.8) | 10 | (23.3) | 24 | (29.6) |
| ≥55 | 5 | (13.2) | 12 | (27.9) | 17 | (21.0) |
| **Marital status** |  |  |  |  |  |  |
| Unmarried | 15 | (39.5) | 14 | (32.6) | 29 | (35.8) |
| Married | 23 | (60.5) | 29 | (67.4) | 52 | (64.2) |
| **Alcohol use** |  |  |  |  |  |  |
| Current user | 35 | (92.1) | 39 | (90.7) | 74 | (91.4) |
| No. days drank in the previous month, median (IQR) | 10 | (5–10) | 15 | (15–20) | 15 | (15–20) |
| No. of drinks drank per day in the previous month, median (IQR) | 1 | (1–1) | 2 | (1–2) | 1 | (1–2) |

**Tobacco use**
|  |  |  |  |
| --- | --- | --- | --- |
| Current user | 22 (57.9) | 35 (81.4) | 57 (70.4) |
| No. of days any tobacco products used in the previous month, median (IQR) | 7.5 (3–15) | 25 (25–25) | 10 (10–25) |
| **Anthropometric profile** |  |  |  |
| Height (cm), median (IQR) | 152.3 (142.2–154.9) | 154.9 (152.4–158) | 154.9 (144.8–157.5) |
| Weight (kg), median (IQR) | 44 (42–48) | 51 (46–58) | 47 (44–53) |
| **Body mass index (kg/m<sup>2</sup>)** |  |  |  |
| Median (IQR) | 20.4 (18.3–22.2) | 21.8 (20–23.3) | 21.1 (19.2–22.9) |
| Under/overweight (<18.5 or ≥25) | 15 (39.5) | 6 (14.0) | 21 (25.9) |
| Normal (18.5–24.9) | 23 (60.5) | 37 (86.0) | 60 (74.1) |
| **Blood pressure (mm of Hg)** |  |  |  |
| SBP, median (IQR) | 122.5 (118.0–132.0) | 135.0 (128.0–153.0) | 130.0 (121.0–137.0) |
| DBP, median (IQR) | 75.0 (70.0–83.0) | 86.0 (79.0–90.0) | 83.0 (74.0–87.0) |
| MAP, median (IQR) | 92.3 (87.0–97.0) | 103.0 (97.0–107.7) | 97.0 (90.3–103.7) |

Abbreviations: DBP, diastolic blood pressure; IQR, interquartile range; MAP, mean arterial pressure; No., number; SBP, systolic blood pressure.

High proportions of males (90.7%) and females (92.1%) currently consume alcohol. In the age group 15–24 years, more than 68% drank alcohol in the past 30 days (data not shown). Overall, more than 2/3^rd^ of the participants currently uses tobacco products (81.4% males, 57.9% females). Approximately 42% of participants in the age group 15–24 years used some form of tobacco product in the preceding 30 days. The preference for tobacco products differed by gender; 87.5% of smokers were females, compared to 97.1% of smokeless tobacco users who were males.

Among females, the median weight was 44 kg (IQR: 42–48), and the median height was 152.3 cm (IQR: 142.2–154.9). Among males, the median weight was 51 kg (IQR: 46–58), and the median height was 154.9 cm (IQR: 152.4–158.0). The Median BMI for the population was 21.1 (IQR: 19.2–22.9) kg/m^2^. About 17% of the participants were underweight (BMI <18.5 kg/m^2^), and 8.6% were overweight (BMI 25 to <30 kg/m^2^). None of the participants were found obese (BMI_≥_30 kg/m^2^).

For females, the medians for systolic blood pressure (SBP) and diastolic blood pressure (DBP) were 122.5 (IQR: 118–132) and 75 (IQR: 70–83) mm Hg, respectively. The same for males were 135 (IQR: 128–153) and 86 (IQR: 79–90) mm of Hg, respectively. Medians for SBP, DBP, and MAP were all higher among males than females across all age groups (Table 1). We found extremely high recordings for SBPs and DBPs among a few participants aged 20 to 30. (Figure 2, A and B). With that exception, in both sexes, the SBP increased steadily with age (Figure 2, A). In males, DBP and MAP both increased significantly with age until the age of 60, when the former began to decline (Figure 2, B and C). In females, DBP and MAP both decreased in early adulthood, followed by a moderate increase until the age of 60, after which the former started to decline.

**Figure 2.**
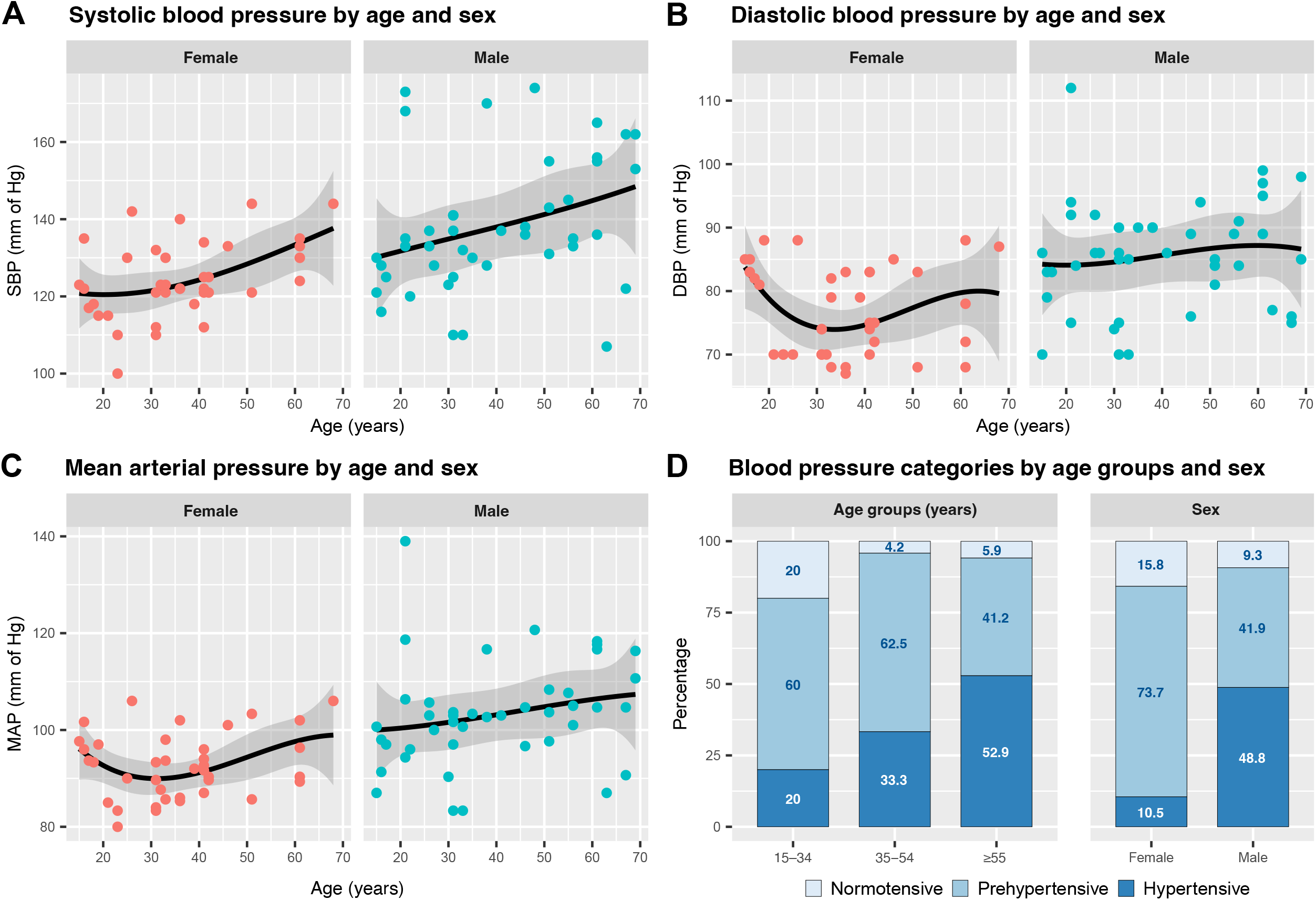
Age-related changes in systolic (A), diastolic (B) and mean arterial (C) blood pressure in nomadic Raute adults. Males are green dots; females are red dots. Displayed curves are third-order polynomial fits. The shaded area represents the 95% confidence interval. The prevalence of hypertension (D) by age group and sex. Bars represent the percentage for each category of normotensive, prehypertensive, and hypertensive participants. The cutoffs for normotension, pre-hypertension, and hypertension were mean SBPs of <120, 120–139, and _≥_140mm of Hg respectively, and/or mean DBPs of <80, 80-89, and _≥_ 90 mm of Hg respectively. Abbreviations: SBP, systolic blood pressure; DBP, diastolic blood pressure; MAP, mean arterial pressure.

#### Prevalence, associated factors, awareness, and treatment of hypertension

The prevalence of hypertension was 10.5% among females, 48.8% among males, and 30.9% overall. Males (48.8%) had a 4.6-fold higher prevalence of hypertension than females (10.5%). Nonetheless, a significant proportion (73.7%) of females were in the pre-hypertensive group (Figure 2, D).

Table 2 summarizes the participants’ sociodemographic characteristics, behavioral risks, and anthropometric profiles by hypertension status. When compared across age groups, the prevalence of hypertension was 2.6 times higher among those 55 and older (52.9%) than among those under 35 (20%). Hypertension was twice more common among those who currently drink (32.4%) than those who do not (14.3%, data not shown). The median days the participants drank alcohol in the previous month were higher among hypertensive than non-hypertensive participants (15 vs. 10 days). Likewise, hypertension was 1.5 times more common among the current tobacco user than the current non-user (30.5 vs. 20.8%). Similarly, the median days of tobacco products used in the previous month among the hypertensive and the non-hypertensive participants were also different (25 vs. 15 days). The median BMIs were almost similar among participants with or without hypertension (21.5 vs. 20.5 Kg/m^2^).

**Table 2.** Socio-demographic, behavioral, and anthropometric factors associated with hypertension among the nomadic Raute population.

| Variables | Non-hypertensive (n=56) |  | Hypertensive (n=25) |  |
| --- | --- | --- | --- | --- |
|  | n | (n%) | n | (n%) |
| **Age (years)** |  |  |  |  |
| Median (IQR) | 33 | (23–42) | 48 | (31–61) |
| 15–34 | 32 | (80.0) | 8 | (20.0) |
| 35–54 | 16 | (66.7) | 8 | (33.3) |
| ≥55 | 8 | (47.1) | 9 | (52.9) |
| **Marital status** |  |  |  |  |
| Unmarried | 20 | (69.0) | 9 | (31.0) |
| Married | 36 | (69.2) | 16 | (30.8) |
| **Alcohol consumption** |  |  |  |  |
| Current drinker | 50 | (67.6) | 24 | (32.4) |
| No. days drank in the previous month, median (IQR) | 10 | (7–15) | 15 | (15–20) |
| No. of drinks drank per day in the previous month, median (IQR) | 1 | (1–1) | 2 | (1–3) |
| **Tobacco use** |  |  |  |  |
| Current user | 37 | (64.9) | 20 | (35.1) |
| Median days (IQR) any tobacco | 15 | (5.0–25.0) | 25 | (22.5–25.0) |

products used in the previous month
| **Body mass index (kg/m<sup>2</sup>)** |  |  |  |
| --- | --- | --- | --- |
| Median (IQR) | 20.5 | (18.5–23.0) | 21.5 (19.6–22.9) |
| Under/Overweight (<18.5 Or ≥25) | 16 | (76.2) | 5 (23.8) |
| Normal (18.5–24.9) | 40 | (66.7) | 20 (33.3) |

Abbreviations: IQR, interquartile range; No., number.

Among the respondents who had hypertension, only 8% were aware of it. None of the hypertensive participants ever received any treatment.

### Qualitative results

A total of 15 Raute participants (10 males, five females, and 33.3% hypertensives) participated in IDIs, with their ages ranging from 15 to 72 years. Four non-Raute key informants (1 social scientist, two social workers, and one health worker) of diverse backgrounds working closely with the Raute community participated in the KIIs (Supplemental Table 1). None of the participants approached refused to participate. Four major themes developed from the thematic analysis are presented below, with some representative narratives from the participants with their age group, sex, hypertension status, and participant number in the parenthesis.

***Theme 1: Socioeconomic transition***

The majority of the key informants observed significant socioeconomic changes in the Raute community over the last decade. According to them, the traditional, foraging- and barter-based economy of Raute is now transitioning towards a predominantly cash-based economy. Most Raute participants, regardless of their hypertension status, agreed that their generation-old belief and view towards money is now changing.

> [People of the previous generation did not accept paper money. They exchanged their wooden wares for grains, flour, and other essentials. They thought accepting paper money was a sin and brought bad luck during their hunting. These days, even our chieftains prefer paper money so that they can purchase goods whenever needed and do not have to carry heavy loads of goods while migrating.] (Raute male, age range 40–45 years, non-hypertensive, IDI#11)

Currently, from the federal and provincial governments combined, each individual, including children from the Raute community, receives a sum of 5,000 Nepali Rupees (Approx. 42 USD) per month.[30] The allowance provided by the government was noted by most participants as the main driver of the socio-economic transition of the community.

> [In the past, we used to earn our living with the use of our muscles. These days we get an allowance. These days we have ease.] (Raute male, age range 55–60 years, hypertensive, IDI#14)
>
> [Previously, my parents must have had so much hardship raising me. We did not have an allowance. They used to make wooden bowls and boxes, go to the village, and exchange them for grains. They ate a little and gave it to me. That’s how they raised me. Nowadays, it is much better.] (Raute male, age range 30-35 years, non-hypertensive, IDI#8)

One of our KII participants indicated that the Rautes’ proximity to settled communities has increased. They now prefer to camp close to market areas where commodities may be easily obtained. Even the Rautes have perceived less of a need to work as the allowance money they receive can be used to purchase food and other essentials.

> [We don’t work as much as before. We can purchase our grains and essentials with an allowance. We go to the forest only for firewood and wood for making our hut. We don’t carry big logs to make wooden wares as much as before.] (Raute male, age range 55–60, hypertensive, IDI#14)

Most of the participants of IDI had witnessed a significant decline in the demand and, therefore, the production of traditional wooden wares. Even though most males identified ‘crafting and trading’ as their primary occupation in our quantitative study, many stated that they now craft wooden wares just to keep the tradition alive. A few participants noted that the change in the livelihood of the Raute was inevitable as modern industrial wares became popular.

> [People don’t like using traditional wooden wares anymore. They prefer to use steel and other metalware these days.] (Raute female, age range 50–55 years, non-hypertensive, IDI#12)

***Theme 2: Change in patterns of alcohol and tobacco use***

### 2.1 Alcohol consumption

Although drinking homemade alcohol has long been a Raute tradition, most participants have noted a shift in alcohol consumption in recent years. Raute, who used to drink traditionally made alcohol occasionally, has lately begun purchasing commercial alcohol from surrounding communities and marketplaces. Many IDIs and KIIs have speculated that their increasing interaction with outsiders could be one of the factors contributing to the Raute community’s rising alcohol usage.

> [We have been drinking homemade alcohol made in wooden vessels since the time of our forefathers. However, the local liquor made and sold by the settled communities and bottled alcohol is getting increasingly popular in recent years. Rautes learned it from the outsiders.] (Raute male, age range 55– 60 years, hypertensive, current drinker, IDI#14)

All the participants of KII reported widespread use of alcohol among Rautes, including youths and children. As evident in our quantitative study, an alarmingly high proportion of Raute youth currently consumes alcohol and tobacco products. One adolescent participant stated that drinking alcohol is common among young Rautes, including children, and they learn it from their elders.

> [These days, big people, even our chieftains, everyone drinks alcohol. So, we, too, drink alcohol. Even small children, smaller than me, consume alcohol. Rautes are like this.] (Raute male, age range 15–20) years, non-hypertensive, current drinker, IDI#1)

One of our KII participants speculated that the provisions of allowance by the government could have an important influence on the recent change in alcohol consumption among the Raute community.

> [Ever since the government started providing them with an allowance, they seem confused about investing this money. They started spending money on alcohol and tobacco.] (Social Scientist, male, age range 50–55 years, KII#3)

However, some Rautes claimed that only a negligible portion of the allowance is spent on buying alcohol.

> [We don’t spend too much money on alcohol. We first buy rice, salt, and flour. We buy alcohol from what remains.] (Raute male, age range 30–35 years, hypertensive, current drinker, IDI#9)

Most of the IDI participants could not clearly outline the adverse health effects of alcohol regardless of their hypertension status. Some participants had certain ideas about the influence of alcohol on their socioeconomic life, general health, and injuries, e.g., falls and burns.

> [We don’t go hunting these days as much as we used to. People nowadays drink alcohol from the allowance they receive. They can’t go hunting when they are drunk. Alcohol makes them feel weak.] (Raute male, age range 15–20 years, non-hypertensive, current drinker, IDI#1)

> [I have heard that alcohol affects our chest and intestine.] (Raute male, age range 15–20 years, hypertensive, current drinker, IDI#3)
>
> [Both I and my wife drink alcohol. Once, we drank and slept near the fireplace. I woke up with a burnt face the next morning. I was so drunk that I did not realize it.] (Raute male, age range 30–35 years, hypertensive, current drinker, IDI#9)

According to a health worker (KII) who works closely with the Raute community, the usage of commercial alcohol has adversely affected the health of Raute youths.

> [Commercial, low-quality alcohol has adversely affected the health of the youth of this community. I saw some young people with jaundice. It must have been caused by alcohol; don’t you think so?] (Health worker, male, age range 25–30 years, KII#1)

### 2.2 Tobacco Use

Most of the Rautes reported using tobacco—the preference for the type of tobacco varied between men and women. We did not find significant differences in patterns, preferences, and perceptions regarding tobacco use among hypertensive and non-hypertensive participants.

> [We, men, prefer chewing tobacco while women prefer cigarettes, hookah, and other smoking tobacco.] (Raute male, age range 55–60 years, hypertensive, current tobacco user, IDI#14)

A recent shift in the pattern with increased use of commercially available tobacco products has been reported by the Rautes.

> [Women used to take hookah in the past. They used to make their own tobacco from tobacco leaves. Nowadays, they can easily buy cigarettes from the marketplace and smoke.] (Raute male, age range 25–30 years, non-hypertensive, IDI#7)

During our IDI, a few Raute asked us for chewing-tobacco. Some participants demanded tobacco products as incentives for participating in our interviews.

> [Raute cannot speak without tobacco. We will talk to you only if you give us tobacco.] (Raute male, 25–30 years, non-hypertensive, current tobacco user, IDI#6)

Most of the respondents were not aware of the adverse effects of tobacco. Only a few participants were able to state that smoking negatively affects health, but they were unable to outline the direct health effects of smoking.

> [Smoking blackens our lungs.] (Raute male, age range 15–20 years, hypertensive, current tobacco user, IDI#3)

***Theme 3: Changing diet and food security***

Most Raute participants had witnessed a significant change in their dietary patterns in recent years. The majority also noted a substantial improvement in food security in recent years compared to the past.

> [In the past, if we could sell our woodenware, we got to eat food; else, we had to either sleep on an empty stomach or borrow food from relatives. These days, we don’t need to sleep hungry.] (Raute male, age range 55–60 years, non-hypertensive, IDI#14)

Historically, the Raute diet consisted of grains obtained through barter and roots, tubers, and herbs collected from the forest, supplemented with bushmeat, according to most participants. The current Raute diet consists chiefly of rice grain and a few vegetables obtained directly from the market or donated by various governmental and non-governmental organizations. Participants reported that hunted game, wild roots, and tubers are less common in their current diet than in the past. The participants did not differ in their perception of changing dietary patterns based on their hypertension status.

> [We previously used to bring air potato, yam, and five-leaf yam from the jungle and eat them. These days, we eat rice all the time.] (Raute male, age range 30–35 years, hypertensive, IDI#9)

Many Raute participants reported that they used to self-process and prepare their food from raw grains. Packaged and processed modern foods have been reported to find their way into the Raute diet in recent years.

> [Back then, we used to go to a watermill or grind it in a large mortar to make flour out of millet and maize to make flatbread. We used to eat roasted maize. Nowadays, we can easily buy Rice and processed wheat flour from the marketplace.] (Raute female, age range 30–35 years, non-hypertensive, IDI#10)
>
> [In our times, we used to eat bayberries, barberries, guava, and various other wild fruits. Young people these days have learned to eat noodles and biscuits.] (Raute male, age range 55–60 years, hypertensive, IDI#14)

***Theme 4: Traditional healthcare practices***

In their ill health, the majority of participants of IDIs reported using various kinds of medicinal herbs based on their indigenous knowledge and seeking traditional healers within their community who practice spiritual healing and herbal medicine. A few participants also stated that they should visit healthcare workers if the illness does not improve with traditional healing, indicating growing awareness of modern healthcare.

> [If we get sick, we try home remedies such as medicinal herbs from the jungle. We also go to the traditional healer to blow charms and assess pulse. If we don’t get better, nowadays they (Outsiders) say we should go to the doctor.] (Raute male, age range 55–60 years, hypertensive, IDI#14)

## Discussion

To the best of our knowledge, this is the first study to assess the status of cardiometabolic health and investigate hypertension as an emerging health concern among endangered Raute hunter-gatherers of Nepal. Overall, 30.9% of the population (10.5% of females and 48.8% of males) were found to have hypertension. A significant majority currently consumes alcohol (91.4%) and tobacco (70.4%), with an alarmingly high prevalence among youths (68 and 42%, respectively). Our qualitative analysis showed that the traditional forage- and barter-based Raute economy had been gradually transitioning into a cash-based economy that is heavily reliant on state incentives, leading to a decline in forest-based activities and an increase in the consumption of purchased foods, beverages, and tobacco products with the increased market involvement.

Although there is still a great paucity of research on HG health, extant data show that HGs and other small-scale subsistence-based populations have a significantly lower prevalence of lifestyle-related diseases, including hypertension.[7,31] HGs with relatively traditional living, such as the Pygmies of Southern Cameroon and the Hadza of Northern Tanzania, for example, were found to have remarkably lower rates of hypertension (3.3 and 13%, respectively).[31–33] Similarly, the Tsimane forager-horticulturists of the Bolivian Amazon also showed a low prevalence of hypertension (2.9%) and no significant increase in BP with age.[34] In contrast to these studies, we found a much higher (30.9%) prevalence of hypertension among nomadic Raute HGs exceeding that of general Nepalese populations (Rural: 19.1–23.8, Urban: 22.4– 25.2%) based on data from the most recent composite national surveys.[18,35] Metanalyses have reported slightly higher national pooled prevalence ranging from 27.3% to up to 32%.[36,37] A recent survey that included samples from former nomadic HGs, including sedentary Rautes from the far west, reported a relatively lower prevalence (23.8%) of hypertension among the indigenous Nepalese population.[38] Among Negritos, Batek, and Jehai HGs from Peninsular Malaysia who receive significant government support and have stable livelihoods demonstrated a much higher (>40%) prevalence of hypertension than our study population.[39,40] The differences in hypertension prevalence observed across these populations might be due to various variables. Aside from genetic, ethnic, and environmental variations, lifestyle, diet, level of physical activity, behavioral risks, and degree of acculturation should all be considered.

The effect of gender and age on BP is well established.[41,42] In the general population, both SBP and DBP rise steadily in early adulthood up to age 50 years, following which SBP steeply rises while DBP markedly declines.[42] In our study, SBP increased steadily with age, whereas DBP did not demonstrate a significant rise. In contrast, Tsimane and Hadza HGs exhibited a less sustained change in BP with the age profiles.[31,43] Across all age categories in our study, males had considerably higher SBP and DBP than females, albeit the difference in BP narrowed slightly after the age of 60 years. This is in agreement with the findings among the general population.[41,44] The BP amplification from central to peripheral arteries increases with body height and is thus more pronounced in males.[45] This may partly explain why brachial artery BP is lower among premenopausal females than among age-matched males. Nonetheless, the origin of sexual dimorphism in BP might be multifaceted, encompassing the combined influence of hormonal, chromosomal, genetic, and morphological variations in body size and socioeconomic and environmental covariates.[46–48] Surprisingly, recent sex-specific analyses indicated that females, compared to males, exhibit a steeper rise in BP with age starting early in life.[46] Among subsistent populations, Tsimane and Pygmy females demonstrated similar gender-related variations in BP.[31,32] Hadza, on the other hand, exhibited only age-related, not gender-related, changes in BP.[33] In our study, the proportion of males with hypertension was significantly higher than that of females. Findings from Tsimane and Pygmies resonate with this.[31,32] In contrast, Hadza females had a higher prevalence of hypertension compared to males.[33]

A significant majority of the participants in our study were current drinkers (91.4%) and some forms of current tobacco users (70.4%). This exceeds the rate of current alcohol (20.8%) and tobacco (28.9%) use among the general Nepalese population.[18] High rates of alcohol (68%) and tobacco (42%) use among the young Raute population under 25 were particularly concerning. Raute youths claim to have learned it from their elders. However, Raute elders believe that their increased interaction with the *‘Duniya’* (the outside world) has influenced the new generation’s heavy use of commercial alcohol and tobacco products. Among Rautes, hypertension was more than twice as common among individuals who currently drink or use tobacco products than those who do not. Interestingly, conventional risk factors, such as alcohol and tobacco use, were not found to be strongly associated with hypertension in many traditional populations, despite their high prevalence.[31,32,38,49,50] This might imply that these populations could have other important hypertension risk factors not caught by existing survey instruments or that they have several protective factors that negate the risk associated with these conventional risk factors, emphasizing the importance of a context-specific approach.

Globally, HGs communities are renowned for their remarkably low obesity.[7] Only a small minority (8.6%) of our study population was overweight (BMI 25–29.9 kg/m^2^), and we found no obesity (BMI _≥_30 kg/m^2^). Among the well-studied HG populations, the prevalence of overweight ranged from 2% among the Hazda to 15–21% among the Tsimane.[7,51] BMI positively correlates with BP among Tsimane and Pygmies.[31,32,52] However, we did not find a correlation between BMI and BP among our study population.

In meta-analyses of several RCTs, the paleolithic diets based on lean meat, fish, fruits, vegetables, root vegetables, eggs, and nuts were shown to have more significant improvements in cardiometabolic parameters such as BP, BMI, lipid profile, and blood sugar level than the guideline-based control diets. However, the benefits were transient, and the strength of the evidence was low.[53,13,54] A transition from the traditional HG diet to a grain-based western diet has been reported to result in a deterioration in general health and an increase in obesity, diabetes, and other metabolic diseases among Australian HGs.[14,55] Interestingly, temporary reversal to the traditional diet among Australian aborigines was further shown to result in marked improvement in carbohydrate and lipid metabolism.[56] Although we did not collect exclusive quantitative data on the Raute diet and nutrition, our qualitative data showed a transition from a traditional forest-based nomadic diet to a grain-based diet with an increasing proportion of market-bought, packaged/processed foods and beverages gradually getting their way into the Raute food culture in recent years. The factors we found, which are also supported by the previous studies, such as depleting forest resources, shrinking traditional barter system, stable cash flow in the form of state incentives, and increasing market involvement, may all be implicated in such dietary transition.[57,58] There is evidence that monetary income and access to the market contributed to a greater intake of energy-dense foods like sugar and oil, as well as a higher rate of obesity and increased drinking and smoking among HGs.[59] Nonetheless, the interaction between market, diet, and HG’s health is complex and warrants more research.

Socioeconomic transition, acculturation, and exposure to Western lifestyles have been equated to an increase in the prevalence of so-called “diseases of civilization” including hypertension, among many traditional communities worldwide.[11,12,60–63] Socioeconomic transition, a major theme of our qualitative analysis, was frequently linked to rising commercial alcohol and tobacco use and changing dietary patterns among Raute. A high rate of alcohol consumption, particularly hazardous consumption, and declining foraging behavior with the socio-economic transition has been well documented among the Congolese BaYaka and Tanzanian Hadza HGs, respectively, which resonate with our findings.[64,65]

The Rautes have rich indigenous knowledge of medicinal herbs. Although traditional healing is still practiced, the Raute is increasingly becoming more aware of modern healthcare. The awareness of the diagnosis among hypertensive participants was 8%, which should not be surprising given the low awareness (∼22%) even among the general population, reflecting overall poor access to and under-utilization of healthcare services in Nepal.

Our study identifies a high burden of hypertension among nomadic Raute HGs in the background of changing socioeconomic status, dietary patterns, and health behaviors. The long-term impact of these changes on Rautes’ health is yet to be explored. From the studies conducted in Australia and South America, it is evident that incidences of cardiovascular and metabolic diseases increase when the population moves away from their traditional lifestyles and gets exposed to modernization.[11,12] The association of various socioeconomic, dietary, and lifestyle factors in populations as simple as HGs emphasizes their potential contribution to the occurrence of cardiovascular disorders like hypertension in other indigenous and general populations in Nepal and elsewhere.

### Study limitation

The several limitations of this study should be noted. The cross-sectional nature of the study limits its ability to establish or refute causal relationships between the variables studied and hypertension. The prevalence of hypertension estimated in this study should be interpreted with caution. The prevalence of hypertension tends to be overestimated in cross-sectional analyses, highlighting the significance of longitudinal monitoring for accurate estimation.[31] We did not collect data on several important factors such as dietary fruits and vegetable consumption, salt intake, level of physical activities, and presence of diabetes mellitus, hyperlipidemia, or central adiposity among our study participants, which would have given deeper insight into the actual CVD risk of this population. Our study was primarily deficit-focused; however, a strengths-based approach highlighting the community’s resilience and strengths alongside its deficiencies would have been more insightful and empowering. We recommend that future research take this critical aspect into account.

Despite these limitations, this study has several strengths. This study is the first to report the prevalence of hypertension and some of its important socio-demographic, behavioral, and metabolic covariates in this unique, difficult-to-reach, endangered population. Enrollment of the entire population, rigorous data collection protocol, and utilization of a mixed-method study to get both quantitative and qualitative perspectives were the strengths of this study.

## Conclusion

We found a high burden of hypertension among nomadic Raute HGs exceeding that of the general Nepalese population. Alcohol and tobacco use was widespread, with alarmingly high rates among youths. The traditional forest-based Raute economy is gradually transitioning to a cash-based economy heavily reliant on government incentives. Consumption of purchased foods, beverages, and tobacco products is increasing in tandem with market involvement. Nonetheless, further research is needed to assess the long-term health impact of these socioeconomic, dietary, and behavioral changes. This study is expected to help appraise concerned policymakers of an emerging health concern and formulate effective context-specific and culturally sensitive community-based interventions to limit the morbidities and mortality associated with hypertension in the endangered nomadic Raute population.

## Data availability statement

All data relevant to the study are included in the article or uploaded as supplementary information. The original datasets are available from the Figshare Digital Repository, https://doi.org/10.6084/m9.figshare.20358837.v2.[66]

## Data Availability

All data relevant to the study are included in the article or uploaded as supplementary information. Datasets are available from the Figshare repository, DOI: [https://doi.org/10.6084/m9.figshare.20358837.v3].

https://doi.org/10.6084/m9.figshare.20358837.v3

## Data Availability

https://doi.org/10.6084/m9.figshare.20358837.v3

## Ethics statements

### Patient consent for publication

Not required.

## Ethical approval

The ethical clearance for this study was obtained from Nepal Health Research Council, Ethical Review Board (Reference No. 2972, Protocol Reg. No. 199/2021P). Informed written consent/assent was obtained from each respondent before the structured questionnaire was administered. Verbal consent was obtained before each IDI and KII and was audio recorded. Participants were informed about the voluntary nature of the study, the maintenance of confidentiality, and the right to refuse or withdraw at any time during the study. No financial incentive was given to the participants of this study.

## Acknowledgment

We are grateful to all our study participants, Raute chieftain Mr. Surya Narayan Shahi and former chieftain Mr. Mahin Bahadur Shahi, for their kind cooperation. We express our sincere gratitude for the facilitation provided by Mr. Binod Kumar BC, field coordinator, Ms. Satya Devi Adhikari, social worker, and the Public Health Service Office, Surkhet. We thank all the data enumerators, and Mr. Ram Prasad Bhandari, for assistance during data collection and processing. We thank Mr. Shital Bhandary (Associate Professor, Patan Academy of Health Sciences, Nepal) for assisting us with statistical analysis. We also acknowledge the language editing and proofreading assistance provided by Dr. Bibiana Cujec (Professor, Faculty of Medicine and Dentistry, University of Alberta, Canada) and Dr. Richard Philip McGuire (Bella Coola General Hospital, British Columbia, Canada).

## Footnotes

### Contributors

TK, UBB, SP, and UP conceptualized the study. TK, UBB, CS, and UP developed the study protocol and contributed to the acquisition of the quantitative data. UBB and TK were involved in acquiring qualitative data. The data was analyzed by TK, SP, CS, and RD, interpreted by all the authors, and visualized by TK and SP. TK, CS, and UP prepared the first draft of the manuscript. RD and MS supervised the study, and reviewed and revised the manuscript for critically important intellectual content. All authors reviewed and approved the final version. TK is responsible for the overall content as guarantor.

### Map disclaimer

The inclusion of any map (including the depiction of any boundaries therein), or of any geographic or locational reference, does not imply the expression of any opinion whatsoever on the part of BMJ concerning the legal status of any country, territory, jurisdiction or area or of its authorities. Any such expression remains solely that of the relevant source and is not endorsed by BMJ. Maps are provided without any warranty of any kind, either express or implied.

### Provenance and peer review

Not commissioned; externally peer reviewed.

## References

1 The Editors of Encyclopaedia Britannica. Nomadism. Britannica. https://www.britannica.com/topic/nomadism (accessed 27 Jun 2021).

2 Lee RB, DeVore I. Man the Hunter: The First Intensive Survey of a Single, Crucial Stage of Human Development— Man’s Once Universal Hunting Way of Life. 1st ed. Routledge 2017. doi:10.4324/9780203786567

3 Asner GP, Knapp DE, Broadbent EN, et al. Selective Logging in the Brazilian Amazon. Science 2005;310:480–2. doi:10.1126/science.1118051

4 Tallavaara M, Seppä H. Did the mid-Holocene environmental changes cause the boom and bust of hunter-gatherer population size in eastern Fennoscandia? The Holocene 2012;22:215–25. doi:10.1177/0959683611414937

5 Armit I, Finlayson B. Hunter-gatherers transformed: the transition to agriculture in northern and western Euope. Antiquity 1992;66:664–76. doi:10.1017/S0003598X00039363

6 Kramer KL, Codding BF. Why forage? hunters and gatherers in the twenty-first century. Albuquerque: : University of New Mexico press 2016.

7 Pontzer H, Wood BM, Raichlen DA. Hunter-gatherers as models in public health. Obes Rev Off J Int Assoc Study Obes 2018;19 Suppl 1:24–35. doi:10.1111/obr.12785

8 Gurven MD, Trumble BC, Stieglitz J, et al. Cardiovascular disease and type 2 diabetes in evolutionary perspective: A critical role for helminths? Evol Med Public Health 2016;2016:338–57. doi:10.1093/emph/eow028

9 Eaton SB, Konner M, Shostak M. Stone agers in the fast lane: Chronic degenerative diseases in evolutionary perspective. Am J Med 1988;84:739–49. doi:10.1016/0002-9343(88)90113-1

10 Lindeberg S. Food and western disease: health and nutrition from an evolutionary perspective. Chichester, U.K.; Ames, Iowa: : Wiley-Blackwell 2010.

11 O’Dea K. Westernisation, insulin resistance and diabetes in Australian Aborigines. Med J Aust 1991;155:258–64. doi:10.5694/j.1326-5377.1991.tb142236.x

12 Steffen PR, Smith TB, Larson M, et al. Acculturation to Western society as a risk factor for high blood pressure: a meta-analytic review. Psychosom Med 2006;68:386–97. doi:10.1097/01.psy.0000221255.48190.32

13 Ghaedi E, Mohammadi M, Mohammadi H, et al. Effects of a Paleolithic Diet on Cardiovascular Disease Risk Factors: A Systematic Review and Meta-Analysis of Randomized Controlled Trials. Adv Nutr 2019;10:634–46. doi:10.1093/advances/nmz007

14 O’Keefe JH, Cordain L. Cardiovascular Disease Resulting From a Diet and Lifestyle at Odds With Our Paleolithic Genome: How to Become a 21st-Century Hunter-Gatherer. Mayo Clin Proc 2004;79:101–8. doi:10.4065/79.1.101

15 Cordain L, Miller JB, Eaton SB, et al. Plant-animal subsistence ratios and macronutrient energy estimations in worldwide hunter-gatherer diets. Am J Clin Nutr 2000;71:682–92. doi:10.1093/ajcn/71.3.682

16 Reinhard J. The Raute: Notes on a Nomadic Hunting and Gathering Tribe of Nepal. Kailash - Journal of Himalayan Studies 1974;2:233–72. doi:10/227326

17 Zhou B, Carrillo-Larco RM, Danaei G, et al. Worldwide trends in hypertension prevalence and progress in treatment and control from 1990 to 2019: a pooled analysis of 1201 population-representative studies with 104 million participants. The Lancet 2021;398:957– 80. doi:10.1016/S0140-6736(21)01330-1

18 Dhimal M BB Bhattarai S, Dixit LP, Hyder MKA, Agrawal N, Rani M, Jha AK. Report on Noncommunicable Risk Factors: STEPS Survey Nepal 2019. Kathmandu: : Nepal Health Research Council 2020. https://www.who.int/docs/default-source/nepal-documents/ncds/ncd-steps-survey-2019-compressed.pdf?sfvrsn=807bc4c6

19 Mesenburg MA, Restrepo-Mendez MC, Amigo H, et al. Ethnic group inequalities in coverage with reproductive, maternal and child health interventions: cross-sectional analyses of national surveys in 16 Latin American and Caribbean countries. Lancet Glob Health 2018;6:e902–13. doi:10.1016/S2214-109X(18)30300-0

20 Adams MW, Sutherland EG, Eckert EL, et al. Leaving no one behind: targeting mobile and migrant populations with health interventions for disease elimination—a descriptive systematic review. BMC Med 2022;20:172. doi:10.1186/s12916-022-02365-6

21 Fortier J. The Ethnography of South Asian Foragers. Annu Rev Anthropol 2009;38:99–114. doi:10.1146/annurev-anthro-091908-164345

22 Shahu MB. Reciprocity practices of nomadic hunter-gatherer R_ā_ute of Nepal. Hunt Gatherer Res 2018;4:257–86.

23 Fortier J. Monkey’s Thigh Is the Shaman’s Meat. In: Kings of the Forest. University of Hawai’i Press 2009. doi:10.21313/hawaii/9780824833220.003.0005

24 Fortier J. Kings of the forest: the cultural resilience of Himalayan hunter-gatherers. Honolulu: : University of Hawai □ i Press 2009.

25 Acharya KP, Paudel PK. Biodiversity in Karnali Province: Current Status and Conservation. Ministry of Industry, Tourism, Forest and Environment, Karnali Province Government, Surkhet, Nepal 2020. http://moitfe.karnali.gov.np/sites/moitfe/files/2020-12/Karnali%20Province%202020_13%20oct%202020_0.pdf

26 Cashin K, Oot L. Guide to Anthropometry: A Practical Tool for Program Planners, Managers, and Implementers. Washington, DC: : Food and Nutrition Technical Assistance III Project (FANTA)/ FHI 360 2018.

27 Central Bureau of Statistics (CBS). Nepal Multiple Indicator Cluster Survey 2019, Survey Findings Report. Kathmandu, Nepal: : Central Bureau of Statistics and UNICEF Nepal 2020. https://www.unicef.org/nepal/reports/multiple-indicator-cluster-survey-final-report-2019

28 Ministry of Health and Population (MOHP) [Nepal] NE ICF International Inc. Nepal Demographic and Health Survey 2016. Kathmandu, Nepal: : Ministry of Health and Population, New ERA, and ICF International, Calverton, Maryland 2017. https://www.dhsprogram.com/pubs/pdf/fr336/fr336.pdf

29 Chobanian AV. The Seventh Report of the Joint National Committee on Prevention, Detection, Evaluation, and Treatment of High Blood PressureThe JNC 7 Report. JAMA 2003;289:2560. doi:10.1001/jama.289.19.2560

30 myRepública. Karnali government to set up ‘Raute Protection Fund’. myRepública 2020.https://myrepublica.nagariknetwork.com/news/karnali-government-to-set-up-raute-protection-fund/

31 Gurven M, Blackwell AD, Rodríguez DE, et al. Does blood pressure inevitably rise with age?: longitudinal evidence among forager-horticulturalists. Hypertens Dallas Tex 1979 2012;60:25–33. doi:10.1161/HYPERTENSIONAHA.111.189100

32 Bika Lele EC, Hermans MP, Bovet P, et al. Prevalence and determinants of blood pressure variability in pygmies of Southern region Cameroon. J Hypertens 2020;38:2198–204. doi:10.1097/HJH.0000000000002529

33 Raichlen DA, Pontzer H, Harris JA, et al. Physical activity patterns and biomarkers of cardiovascular disease risk in hunter-gatherers. Am J Hum Biol 2017;29:e22919. doi:10.1002/ajhb.22919

34 Kaplan H, Thompson RC, Trumble BC, et al. Coronary atherosclerosis in indigenous South American Tsimane: a cross-sectional cohort study. The Lancet 2017;389:1730–9. doi:10.1016/S0140-6736(17)30752-3

35 Das Gupta R, Bin Zaman S, Wagle K, et al. Factors associated with hypertension among adults in Nepal as per the Joint National Committee 7 and 2017 American College of Cardiology/American Heart Association hypertension guidelines: a cross-sectional analysis of the demographic and health survey 2016. BMJ Open 2019;9:e030206. doi:10.1136/bmjopen-2019-030206

36 Huang Y, Guo P, Karmacharya BM, et al. Prevalence of hypertension and prehypertension in Nepal: a systematic review and meta-analysis. Glob Health Res Policy 2019;4:11. doi:10.1186/s41256-019-0102-6

37 Dhungana RR, Pandey AR, Shrestha N. Trends in the Prevalence, Awareness, Treatment, and Control of Hypertension in Nepal between 2000 and 2025: A Systematic Review and Meta-Analysis. Int J Hypertens 2021;2021:e6610649. doi:10.1155/2021/6610649

38 Denekew TW, Gautam Y, Bhandari D, et al. Prevalence and determinants of hypertension in underrepresented indigenous populations of Nepal. PLOS Glob Public Health 2022;2:e0000133. doi:10.1371/journal.pgph.0000133

39 Mokhsin A, Mokhtar SS, Mohd Ismail A, et al. Observational study of the status of coronary risk biomarkers among Negritos with metabolic syndrome in the east coast of Malaysia. BMJ Open 2018;8:e021580. doi:10.1136/bmjopen-2018-021580

40 Phipps ME, Chan KK, Naidu R, et al. Cardio-metabolic health risks in indigenous populations of Southeast Asia and the influence of urbanization. BMC Public Health 2015;15:47. doi:10.1186/s12889-015-1384-3

41 Franklin SS, Gustin W, Wong ND, et al. Hemodynamic Patterns of Age-Related Changes in Blood Pressure: The Framingham Heart Study. Circulation 1997;96:308–15. doi:10.1161/01.CIR.96.1.308

42 Cheng S, Xanthakis V, Sullivan LM, et al. Blood Pressure Tracking Over the Adult Life Course: Patterns and Correlates in the Framingham Heart Study. Hypertension 2012;60:1393–9. doi:10.1161/HYPERTENSIONAHA.112.201780

43 Barnicot NA, Bennett FJ, Woodburn JC, et al. Blood pressure and serum cholesterol in the Hadza of Tanzania. Hum Biol 1972;44:87–116.

44 Pearson JD, Morrell CH, Brant LJ, et al. Age-Associated Changes in Blood Pressure in a Longitudinal Study of Healthy Men and Women. J Gerontol A Biol Sci Med Sci 1997;52A:M177–83. doi:10.1093/gerona/52A.3.M177

45 London GM, Guerin AP, Pannier B, et al. Influence of Sex on Arterial Hemodynamics and Blood Pressure: Role of Body Height. Hypertension 1995;26:514–9. doi:10.1161/01.HYP.26.3.514

46 Ji H, Kim A, Ebinger JE, et al. Sex Differences in Blood Pressure Trajectories Over the Life Course. JAMA Cardiol 2020;5:255. doi:10.1001/jamacardio.2019.5306

47 Arnold AP, Cassis LA, Eghbali M, et al. Sex Hormones and Sex Chromosomes Cause Sex Differences in the Development of Cardiovascular Diseases. Arterioscler Thromb Vasc Biol 2017;37:746–56. doi:10.1161/ATVBAHA.116.307301

48 Dickerson JA, Nagaraja HN, Raman SV. Gender-Related Differences in Coronary Artery Dimensions: A Volumetric Analysis. Clin Cardiol 2010;33:E44–9. doi:10.1002/clc.20509

49 Manimunda SP, Sugunan AP, Benegal V, et al. Association of hypertension with risk factors & hypertension related behaviour among the aboriginal Nicobarese tribe living in Car Nicobar Island, India. Indian J Med Res 2011;133:287–93.

50 Pavan L, Casiglia E, Braga LM, et al. Effects of a traditional lifestyle on the cardiovascular risk profile: the Amondava population of the Brazilian Amazon. Comparison with matched African, Italian and Polish populations. J Hypertens 1999;17:749–56. doi:10.1097/00004872-199917060-00005

51 Gurven M, Jaeggi AV, Kaplan H, et al. Physical activity and modernization among Bolivian Amerindians. PloS One 2013;8:e55679. doi:10.1371/journal.pone.0055679

52 Gurven M, Kaplan H, Winking J, et al. Inflammation and Infection Do Not Promote Arterial Aging and Cardiovascular Disease Risk Factors among Lean Horticulturalists. PLoS ONE 2009;4:e6590. doi:10.1371/journal.pone.0006590

53 Manheimer EW, van Zuuren EJ, Fedorowicz Z, et al. Paleolithic nutrition for metabolic syndrome: systematic review and meta-analysis. Am J Clin Nutr 2015;102:922–32. doi:10.3945/ajcn.115.113613

54 Dinu M, Pagliai G, Angelino D, et al. Effects of Popular Diets on Anthropometric and Cardiometabolic Parameters: An Umbrella Review of Meta-Analyses of Randomized Controlled Trials. Adv Nutr 2020;11:815–33. doi:10.1093/advances/nmaa006

55 O’Dea K, Spargo RM, Akerman K. The Effect of Transition from Traditional to Urban Life-Style on the Insulin Secretory Response in Australian Aborigines. Diabetes Care 1980;3:31–7. doi:10.2337/diacare.3.1.31

56 O’dea K. Marked Improvement in Carbohydrate and Lipid Metabolism in Diabetic Australian Aborigines After Temporary Reversion to Traditional Lifestyle. Diabetes 1984;33:596–603. doi:10.2337/diab.33.6.596

57 Gurung OP, Rawal N, Bista PB. Raute of Nepal. First edition. Kathmandu, Nepal: : Central Department of Sociology/Anthropology, Tribhuvan University 2014.

58 Paudel M. Resistance and change.A case study of economic changes and its effect on language, food habits and dress of the nomadic hunting-gathering Raute of Nepal. 2016.https://hdl.handle.net/10037/9783

59 Donders I, Barriocanal C. The Influence of Markets on the Nutrition Transition of Hunter-Gatherers: Lessons from the Western Amazon. Int J Environ Res Public Health 2020;17:6307. doi:10.3390/ijerph17176307

60 DiBello JR, McGarvey ST, Kraft P, et al. Dietary Patterns Are Associated with Metabolic Syndrome in Adult Samoans. J Nutr 2009;139:1933–43. doi:10.3945/jn.109.107888

61 LaMonica LC, McGarvey ST, Rivara AC, et al. Cascades of diabetes and hypertension care in Samoa: Identifying gaps in the diagnosis, treatment, and control continuum – a cross-sectional study. Lancet Reg Health - West Pac 2022;18:100313. doi:10.1016/j.lanwpc.2021.100313

62 Port Lourenço AE, Ventura Santos R, Orellana JDY, et al. Nutrition transition in Amazonia: Obesity and socioeconomic change in the Suruí Indians from Brazil. Am J Hum Biol 2008;20:564–71. doi:10.1002/ajhb.20781

63 Ramachandran A, Snehalatha C. Rising Burden of Obesity in Asia. J Obes 2010;2010:1–8. doi:10.1155/2010/868573

64 Knight JK, Salali GD, Sikka G, et al. Quantifying patterns of alcohol consumption and its effects on health and wellbeing among BaYaka hunter-gatherers: A mixed-methods cross-sectional study. PLOS ONE 2021;16:e0258384. doi:10.1371/journal.pone.0258384

65 Pollom TR, Herlosky KN, Mabulla IA, et al. Changes in Juvenile Foraging Behavior among the Hadza of Tanzania during Early Transition to a Mixed-Subsistence Economy. Hum Nat 2020;31:123–40. doi:10.1007/s12110-020-09364-7

66 [dataset] 66 Koirala Tapendra, BC, Udaya Bahadur, Shrestha, Carmina, et al. Arterial hypertension and its covariates among nomadic Raute hunter-gatherers of Western Nepal: a mixed-method study. Figshare Digit. Repos. 2022. doi:10.6084/M9.FIGSHARE20358837.V2

