## Supplemental Figure 1 for "Arterial hypertension and its covariates among nomadic Raute hunter-gatherers of Western Nepal: a mixed-method study"

Total Raute population: 144

Did not meet inclusion criteria: 59 - Under 15 years of age: 56 - Pregnant woåmen: 3

Eligible population: 85

Included in the final analysis: 81 - Male: 43 - Female: 38

- Declined: 2 - Suspected COVID-19: 1 - Excluded from final analysis: 1 (Extreme of age)

Supplemental Figure 1

The process of participant selection for the quantitative study.
