## Supplemental Table 1 for "Arterial hypertension and its covariates among nomadic Raute hunter-gatherers of Western Nepal: a mixed-method study"

**Supplemental Table 1:** Participant characteristics of the qualitative study

| Interviews | Variables | Categories | Count | Percent |
|---|---|---|---|---|
| In-depth interviews (N=15) | Age | 15–29 | 6 | 40.0 |
| | | 30–44 | 5 | 33.3 |
| | | ≥45 | 4 | 26.7 |
| | Sex | Female | 5 | 33.3 |
| | | Male | 10 | 66.7 |
| | Marital state | Unmarried | 5 | 33.3 |
| | | Married | 10 | 66.7 |
| | Current drinker | Yes | 14 | 93.3 |
| | Current tobacco user | Yes | 12 | 80.0 |
| | Hypertension | No | 10 | 66.7 |
| | | Yes | 5 | 33.3 |
| | Education | No formal education | 15 | 100.0 |
| Key-informant interviews (N=4) | Age | <45 | 2 | 50.0 |
| | | ≥45 | 2 | 50.0 |
| | Sex | Female | 1 | 25.0 |
| | | Male | 3 | 75.0 |
| | Marital state | Married | 4 | 100.0 |
| | Education | No formal education | 1 | 25.0 |
| | | Diploma level | 1 | 25.0 |
| | | Bachelor’s level | 1 | 25.0 |
| | | Masters and higher | 1 | 25.0 |
