## Supplemental File 1 for "Arterial hypertension and its covariates among nomadic Raute hunter-gatherers of Western Nepal: a mixed-method study"

### Supplemental File 1: Tools for the quantitative study

### A. Household questionnaire (English Version)

**Household Number: [	] [	]**

Greetings! My name is ____________________ . We are here to conduct a survey entitled “Prevalence and factors associated with hypertension among Nomadic Rāute population of Western Nepal: a mixed-method study” to know the situation of hypertension among the Rāute community. First, I would like to ask a few questions about your household. The questions usually take about 15 to 20 minutes. Following this, I may ask to conduct additional interviews with you or other individual members of your household aged 15 years or older. The personal data provided by you in this questionnaire will be kept confidential and the details will not be shown, given, or told to any other persons other than the members of our survey team. No part of this interview is being recorded in tape or video. If you do not wish to answer a question or stop the interview, please let me know. Do you have any questions? May I begin the interview now?

| **A** | **B** | **C** | **D** | **E** | **F** | **G** | **H** |
| --- | --- | --- | --- | --- | --- | --- | --- |
| Line No. | Frist, please tell me the name of each person who usually lives here, starting with the head of the household. | What is the relationship of (name) to (name of the head of household)? | Is (name) male or female?  1 FEMALE  2 MALE | How old is (name)?  *Record in completed years.* | What is (name)’s current marital status?  *(if age 10 years or above)* | Circle line number of all women and men 15 years and above. | Is (name) currently at the home?  1 YES  2 NO |
| 01 |  | **1** |  |  |  | 01 |  |
| 02 |  |  |  |  |  | 02 |  |
| 03 |  |  |  |  |  | 03 |  |
| 04 |  |  |  |  |  | 04 |  |
| 05 |  |  |  |  |  | 05 |  |
| 06 |  |  |  |  |  | 06 |  |
| 07 |  |  |  |  |  | 07 |  |
| 08 |  |  |  |  |  | 08 |  |
| 09 |  |  |  |  |  | 09 |  |
| 10 |  |  |  |  |  | 10 |  |

| **Continuation Sheet used?** |  | **Codes for relationship to head of household** | | **Codes for marital status** |  | **Any members not at home?** |
| --- | --- | --- | --- | --- | --- | --- |
| 1 Yes  2 No |  | 01 Head  02 Wife/Husband  03 Son/Daughter  04 Son-in-law/Daughter-in-law  05 Grandchild | 06 Parent  07 Parent-in-law  08 Brother/Sister  09 Other | 1 Never married 2 Married 3 Widowed 4 Divorced 5 Separated |  | 1 Yes 2 No  Female =  Male =  Total = |

### C. Individual questionnaire (English Version)

| **QN** | **Question** | **Possible Response** | **Skip** |
| --- | --- | --- | --- |
| A | Participant ID | [ ] [ ] [ ] |  |
| B | Sex (*Record Female/Male as observed*) | Female 1  Male 2 |  |
| C | What is your completed age in years? | [ ] [ ] (years) |  |
| D | What is your current marital status? | Never married 1 Married 2 Widowed 3  Divorced 4 Separated 5 |  |
| E | Have you ever attended school or any education programme? | Yes 1  No 2 |  |
| F | What is your major occupation? | Unemployed 1  Carving woodenwares 2 Household chores 3 Others (Specify) 4  _________________________ |  |
| G1 | *Is the respondent a married woman of age 15-49 years?* | Yes 1  No 2 | 2→Go to H1 |
| G2 | Are you currently pregnant? | Yes 1  No 2 | 1 → STOP |
| Now I am going to ask you some questions about alcohol consumption. | | | |
| H1 | [Have you ever consumed an alcoholic drink such as beer, wine, or any homemade alcohol drinks?] (USE SHOWCARDS 1a) | Yes 1  No 2 | 2→Go to I1 |
| H2 | During the last one month, on how many days did you have at least one drink of alcohol?  (USE SHOWCARDS 1b)  *Specify the number of days drank. If the respondent did not drink, record ‘00’. If ‘Every day’ or ‘Almost every day’, record ‘30’.* | Number of days drank (*1-29*) [ ] [ ]  Did not drink in last one month 00  Every day / almost every day 30 |  |
| H3 | During the last one month, when you drank alcohol, how many standard drinks on average did you have during one drinking occasion?  (USE SHOWCARDS 1b) | Number of drinks [ ] [ ] |  |
| Next, I would like to ask you some questions about tobacco use. | | | |
| I1 | [Have you ever smoked any tobacco products such as cigarettes, *bidis*, cigars, hookah, or any local/homemade tobacco products?] (USE SHOWCARDS 2a) | Yes 1  No 2 | 2→Go to I3 |
| I2 | During the last month, how many days did you smoke any tobacco products?  *Specify the number of days smoked. If respondent did not smoke, record ‘00’. If ‘Every day’ or ‘Almost every day’, record ‘30’.* | Number of days smoked (*1-29*) [ ] [ ]  Did not smoke in last one month 00  Every day / almost every day 30 |  |
| I3 | [Have you ever used any smokeless tobacco products such as chewing tobacco, snuff, or local/homemade products?] (USE SHOWCARDS 2b) | Yes 1  No 2 | 2→Go to J1 |
| I4 | [During the last one month, on how many days did you use any smokeless tobacco products such as chewing tobacco, snuff, or local/homemade products?]  *Specify the number of days any smokeless tobacco products were used. If respondent did not use any products, record ‘00’. If ‘Every day’ or ‘Almost every day’, record ‘30’.* | Number of days used (*1-29*) [ ] [ ]  Did not use in last one month 00  Every day / almost every day 30 |  |
| J1 | Have you ever been told by doctor or other health worker that you have raised blood pressure, hypertension or pressure? | Yes 1  No 2 | 2→Go to K1 |
| J2 | In the past two weeks, have you taken any medication for raised blood pressure prescribed by a doctor or other health worker? | Yes 1  No 2 |  |
| K1 | Have youever been told by a doctor or other health worker that you have raised blood sugar or diabetes ? | Yes 1  No 2 | 2→Go to L1 |
| K2 | In the past two weeks, have you taken any medication for diabetes, or raised blood sugar prescribed by a doctor or other health worker? | Yes 1  No 2 |  |
| L1 | In the last 14 days, have you experienced ANY of these symptoms?  *(Multiple responses possible)* | Fever 1  Chills/rigour 2  Dry cough 3  Conjunctivitis 4 Shortness of breath 5  Runny or congested nose 6  Headache 7  Diarrhea/loose motion 8  Muscle aches/joint pain 9  Sore throat 10  Nausea/vomiting 11  Loss of taste or smell 12 Others (Specify) 13 _____________  None of the above 00 | 1 to 13 → STOP |
| L2 | Have you been in close contact (within 2 metres/ 6 feet for more than 10 minutes total over 24 hours) in the last 14 days with someone who has tested positive for COVID-19 (with a rapid antigen test or laboratory based test) | Yes 1  No 2  Don’t know 3 | 1 → STOP |
| M | Blood pressure—Systolic (mm of Hg) | [ ][ ][ ] [ ][ ][ ] [ ][ ][ ] |  |
|  |  | Measurement 1 Measurement 2 Measurement 3 |  |
| N | Blood pressure—Diastolic (mm of Hg) | [ ][ ][ ] [ ][ ][ ] [ ][ ][ ] |  |
|  |  | Measurement 1 Measurement 2 Measurement 3 |  |
| O | Weight (kg) | [ ] [ ] [ ] (kg) |  |
| P | Height (cm) | [ ] [ ] [ ] (cm) |  |

### Show Cards

Showcards were adopted from STEPS Survey Nepal 2019. ^[1]^

**Reference**

1.	Dhimal M BB Bhattarai S, Dixit LP, Hyder MKA, Agrawal N, Rani M, Jha AK. Report on Noncommunicable Risk Factors: STEPS Survey Nepal 2019. Kathmandu: Nepal Health Research Council; 2020. Available: https://www.who.int/docs/default-source/nepal-documents/ncds/ncd-steps-survey-2019-compressed.pdf?sfvrsn=807bc4c6
