## Supplemental File 2 for "Arterial hypertension and its covariates among nomadic Raute hunter-gatherers of Western Nepal: a mixed-method study"

### Supplemental File 2: Tools for the qualitative study

#### A. In-depth Interview guide for Raute Participants [English]

| Interview conduction date:  Interviewee: **Rāute Participant**  Name of Interviewer:  Interview start time:  Interview end time:  Place of interview:  Recorded: Yes | | |
| --- | --- | --- |
| Age : [ ] [ ] [ ] years Sex : F / M Hypertension : Yes / No / Don’t know | | |
| Consent | Explain the study and ask for the consent, then record the consent when you turn on the microphone.  *Eg: “Thank you for talking to me today… are you happy to take part in the study?”*  **Turn the recorder on.**  *Ok, so I have turned the microphone on. I just want to ask you again, are you happy to take part in this study by talking with me today?*  Let’s start the interview. | |
| **Themes** | **Questions** | **Probe** |
| Livelihood | What is the main form of your livelihood now?  Have you seen any difference between 10/15 years ago and now? Why? | Differences in terms of:   - Hunting/Foraging - Carving and trading woodenwares - Income generation - Hardship |
| Activity level | In your opinion, what changes have taken place in terms of activities of daily living of the Rāutes in the last 10/15 years? | Differences in terms of:   - Going to the forest, fetching firewood, carrying loads, travel, transportation, etc. |
| Diet | In your opinion, is there any difference between the food/dishes eaten now by the Raute community from what used to be eaten 10/15 years back? | Ask what changes have taken place in the availability and use of traditional forest-based foods (e.g. yam, air potato, vegetables etc.)?  Ask about the use of market food / local food. (Noodles, biscuits, chocolates, hotel food .........)  Why and how did such a change come about? |
|  | What changes have the Raute community experienced in terms of daily food intake, its adequacy, availability, etc., 10/15 years ago and now? |  |
| Alcohol and Tobacco use | In your opinion, is drinking alcohol good or bad for health? Why?  In your opinion, is smoking/chewing tobacco good or bad for health? Why? | If good– why? Explore  If bad– why? Explore |
|  | In your opinion, is there any change in the use of tobacco products (Cigarettes, Chewing tobacco, Hookah,) 10-15 years ago and now in the Raute community? | If so, why such a difference? |
|  | In your opinion, is there any change in the use of alcoholic beverages (Beer, wine, vodka, homemade alcohol products etc.) 0-15 years ago and now in the Raute community? | If so, why such a difference? |
|  | In your opinion, in terms of alcohol consumption and tobacco use, is there any difference between men and women of Raute community? | If so, what are the differences? |
| Concept of ill-health & Knowledge of NCDs | Now we are near the end of the interview.  In your opinion, why do people get sickness or illness? | Ask, what do they do when they get sick? |
|  | Have you heard of non-communicable diseases? Example? | If says ‘No’ → Probe   - High blood pressure (high BP), diabetes (*Raised blood sugar)*, COPD (*Chronic lung disease)*, Cancer etc. |
|  | Have you heard of hypertension/high blood pressure? |  |
|  | What do you think causes high blood pressure (high BP) in humans? | Probe:  Smoking, Alcohol, Obesity, Stress, Salt intake, Oil intake, Dietary fibers etc. |
| Thank you very much for participating in this study.  END | | |

#### B. In-depth Interview guide for non-Raute Participants [English]

| Interview conduction date:  Interviewee: **non**-**Rāute Participant**  Name of Interviewer:  Interview start time:  Interview end time:  Place of interview:  Recorded: Yes | | |
| --- | --- | --- |
| Age : [ ] [ ] [ ] years Sex : F / M Designation: | | |
| Consent | Explain the study and ask for the consent, then record the consent when you turn on the microphone.  *Eg: “Thank you for talking to me today… are you happy to take part in the study?”*  **Turn the recorder on.**  *Ok, so I have turned the microphone on. I just want to ask you again, are you happy to take part in this study by talking with me today?*  Let’s start the interview. | |
| **Themes** | **Questions** | **Probe** |
| Livelihood | What is the main form of Raute’s livelihood now?  Have you seen any difference between 10/15 years ago and now? Why? | Differences in terms of:   - Hunting/Foraging - Carving and trading woodenwares - Income generation - Hardship |
| Activity level | In your opinion, what changes have taken place in terms of activities of daily living of the Rāutes in the last 10/15 years? | Differences in terms of:   - Going to the forest, fetching firewood, carrying loads, travel, transportation, etc. |
| Diet | In your opinion, is there any difference between the food/dishes eaten now by the Raute community from what used to be eaten 10/15 years back? | Ask what changes have taken place in the availability and use of traditional forest-based foods (e.g., yam, air potato, vegetables etc.)?  Ask about the use of market food / local food. (Noodles, biscuits, chocolates, hotel food .........)  Why and how did such a change come about? |
|  | What changes have the Raute community experienced in terms of daily food intake, its adequacy, availability, etc., 10/15 years ago and now? |  |
| Alcohol and Tobacco use | In your opinion, is there any change in the use of tobacco products (Cigarettes, Chewing tobacco, Hookah,) 10-15 years ago and now in the Raute community? | If so, why such a difference? |
|  | In your opinion, is there any change in the use of alcoholic beverages (Beer, wine, vodka, homemade alcohol products etc.) 0-15 years ago and now in the Raute community? | If so, why such a difference? |
|  | In your opinion, in terms of alcohol consumption and tobacco use, is there any difference between men and women of Raute community? | If so, what are the differences? |
| Thank you very much for participating in this study. | | |
| END | | |
