## Supplemental File 3 for "Arterial hypertension and its covariates among nomadic Raute hunter-gatherers of Western Nepal: a mixed-method study"

**Operational definitions**

- **Current smokers** were those who self-reported smoking tobacco products within the last 30 days.
- **Current alcohol drinkers** were those who self-reported consuming alcohol within the last 30 days.
- **Body mass index (BMI)** was categorized as underweight (BMI <18.5 kg/m^2^), normal (BMI 18.0–24.9 kg/m^2^), overweight (BMI 25.0–29.9 kg/m^2^), or obese (BMI ≥30 kg/m^2^).
- The blood pressure (BP) was categorized based on the JNC 7 recommendations.[29]
  - **Hypertension** was defined as systolic blood pressure (SBP) ≥ 140 mm Hg and/or diastolic blood pressure (DBP) ≥ 90 mm Hg or the current use of antihypertensive medication irrespective of the blood pressure during the survey.
  - **Prehypertension** was defined as SBP of 120–139 mm Hg and/or DBP of 80–89 mm Hg. Awareness of hypertension was defined as a self-reported condition of hypertension that a doctor or other health worker had diagnosed prior to the data collection.
  - **Treatment of hypertension** was defined as self-reported use of any antihypertensive medication in the last two weeks before the interview.
